# Outcomes of a Multidisciplinary Approach to Management of Mavacamten in Obstructive Hypertrophic Cardiomyopathy

**DOI:** 10.1101/2024.03.05.24303837

**Authors:** Hanna Jensen, Zaid Yousif, Trina Huynh, Megan Kraushaar, Marcy Reed, Trista Boarman, Jorge Silva Enciso, Andrew Willeford

## Abstract

**Background:** Traditional treatments for obstructive hypertrophic cardiomyopathy (oHCM) include beta-blockers, calcium channel blockers, and disopyramide. Mavacamten, a novel cardiac myosin inhibitor, is a promising oHCM therapy but has practical challenges limiting its use. This study aimed to describe a clinic workflow for mavacamten management in a real-world setting, addressing challenges such as cost, drug interactions, and monitoring requirements. The focus was on reducing patient-level costs while ensuring feasibility and efficiency.

**Methods:** A retrospective analysis was conducted on 34 oHCM patients considered for mavacamten between May 2022 and May 2023. The clinic workflow involved cardiologist assessment, pharmacist evaluation of drug interactions, enrollment in the mavacamten REMS program, cost reduction measures, and initiation monitoring through scheduled echocardiograms. An algorithm detailing steps and tools used in this workflow is provided.

**Results:** Of the 34 patients, 21 (62%) were initiated on mavacamten and followed for up to 1 year on therapy. Cost assessments indicated reduced out-of-pocket expenses with assistance programs. The median time from referral to first fill was 22 days. Patients demonstrated high adherence (99.1%) measured by proportion of days covered. Echocardiogram follow-ups showed significant improvements in left ventricular outflow tract parameters with no patients having a decrease in left ventricular ejection fraction to less than 50%.

**Conclusions:** The described workflow effectively addressed challenges associated with mavacamten management, emphasizing roles for clinic personnel, cost reduction strategies, and structured patient monitoring. While the workflow’s specifics may need adaptation in different settings, this report provides valuable insights for clinics implementing structured mavacamten management approaches.

## INTRODUCTION

Hypertrophic cardiomyopathy (HCM) is a genetic myocardial disease with an estimated prevalence of 1 case per 500 persons and is defined by cardiac hypertrophy with variable phenotypic expression.^1^ Hypertrophy predominantly occurs in the basal interventricular septum but may occasionally occur in the apex.^2^ Pathophysiological characteristics include hypercontractility, diastolic dysfunction, and dynamic outflow tract obstruction which is found in approximately two thirds of patients.^3,4^ In obstructive hypertrophic cardiomyopathy (oHCM), the obstruction is considered the primary driver of symptoms which may include dyspnea, chest pain, palpitations, and syncope.^5^ Complications include heart failure, atrial fibrillation, and sudden cardiac death.^6,7,8^

Historically, treatment options consisted of beta-blockers, non-dihydropyridine calcium channel blockers, and in refractory cases, disopyramide—a class 1A anti-arrhythmic.^9,10^ These treatments are considered to decrease the outflow tract obstruction through their negative inotropic effects, leading to symptomatic relief. In April of 2022, the United States Food and Drug Administration approved a new medication for oHCM, the novel cardiac myosin inhibitor, mavacamten.^11^ Mavacamten reversibly inhibits cardiac myosin, effectively reducing the number of myosin-actin cross bridges that can form during the contraction and relaxation cycle of the cardiac sarcomere.^12^ Resultant effects include reduction in left ventricular outflow tract (LVOT) obstruction, improved cardiac filling pressure, improved energy consumption, and attenuated cardiac contractility.

Recent findings from the EXPLORER-HCM and VALOR-HCM trials demonstrated mavacamten as a promising treatment option.^13,14^ The studies showed decreased symptom burden, increased functional capacity, and reduced need for septal reduction therapy (SRT) compared to placebo. Importantly, a small number of patients were found to have a reduction in left ventricular ejection fraction (LVEF) to less than 50% that was reversible after holding therapy.^13^ This finding is responsible for implementation of the mavacamten Risk Evaluation and Mitigation Strategy (REMS) program which aims to detect heart failure secondary to systolic dysfunction through screening with regular echocardiograms.^11^ Furthermore, the program intends to screen for drug-drug interactions that may decrease mavacamten metabolism and subsequently put patients at increased risk for systolic dysfunction.

Mavacamten has significant costs to patients, and the management is complex. It requires frequent echocardiogram monitoring, dose adjustments based on Valsalva left ventricular outflow tract (VLVOT) gradient changes, and extensive screening for drug interactions.^11^ These requirements pose significant challenges which emphasize the need for a personalized clinic workflow to optimize the initiation and management of patients on mavacamten. This study will describe the clinic workflow at a hypertrophic cardiomyopathy center of excellence which is aimed at reducing patient-level costs and establishing feasibility of mavacamten management in a real-world setting.

## METHODS

### Study Population

This was a single-center retrospective, descriptive analysis of patients with a history of oHCM considered for mavacamten at a hypertrophic cardiomyopathy center of excellence between May 1, 2022, and May 31, 2023. This study was approved by the UC San Diego Health institutional review board. Informed consent was not required. All patients were cared for using a workflow that was implemented by the study investigators in May of 2022. The workflow was created to lower patient-level costs and ensure clinic efficiency regarding the initiation and follow-up of mavacamten. Roles in the workflow were assigned for cardiologists, pharmacists, nurses, and echocardiogram coordinators. Pharmacists had a collaborative practice agreement with the cardiologists which allowed participation in drug therapy management.

### Workflow description

The workflow is presented in Figure 1. Patients are first seen by a cardiologist who determines initial eligibility for therapy based on symptom assessment and recent echocardiography parameters (i.e., LVEF ≥55%). Eligible patients are then referred to the pharmacist who assesses drug interactions and the need for contraception. Mavacamten is primarily metabolized by cytochrome P450 (CYP) 3A4 and CYP 2C19 and is contraindicated in patients taking a moderate to strong CYP 2C19 inhibitor, strong CYP 3A4 inhibitor, moderate to strong CYP 2C19 inducer, or moderate to strong CYP 3A4 inducer.^15^ Furthermore, mavacamten is an inducer of CYP 3A4, CYP 2C9, and CYP 2C19, which requires assessment of interactions from mavacamten on other medications including hormonal contraceptives. Effective contraception is needed in patients of childbearing potential since in utero exposure to mavacamten may cause fetal harm.^15^

**Figure 1.**
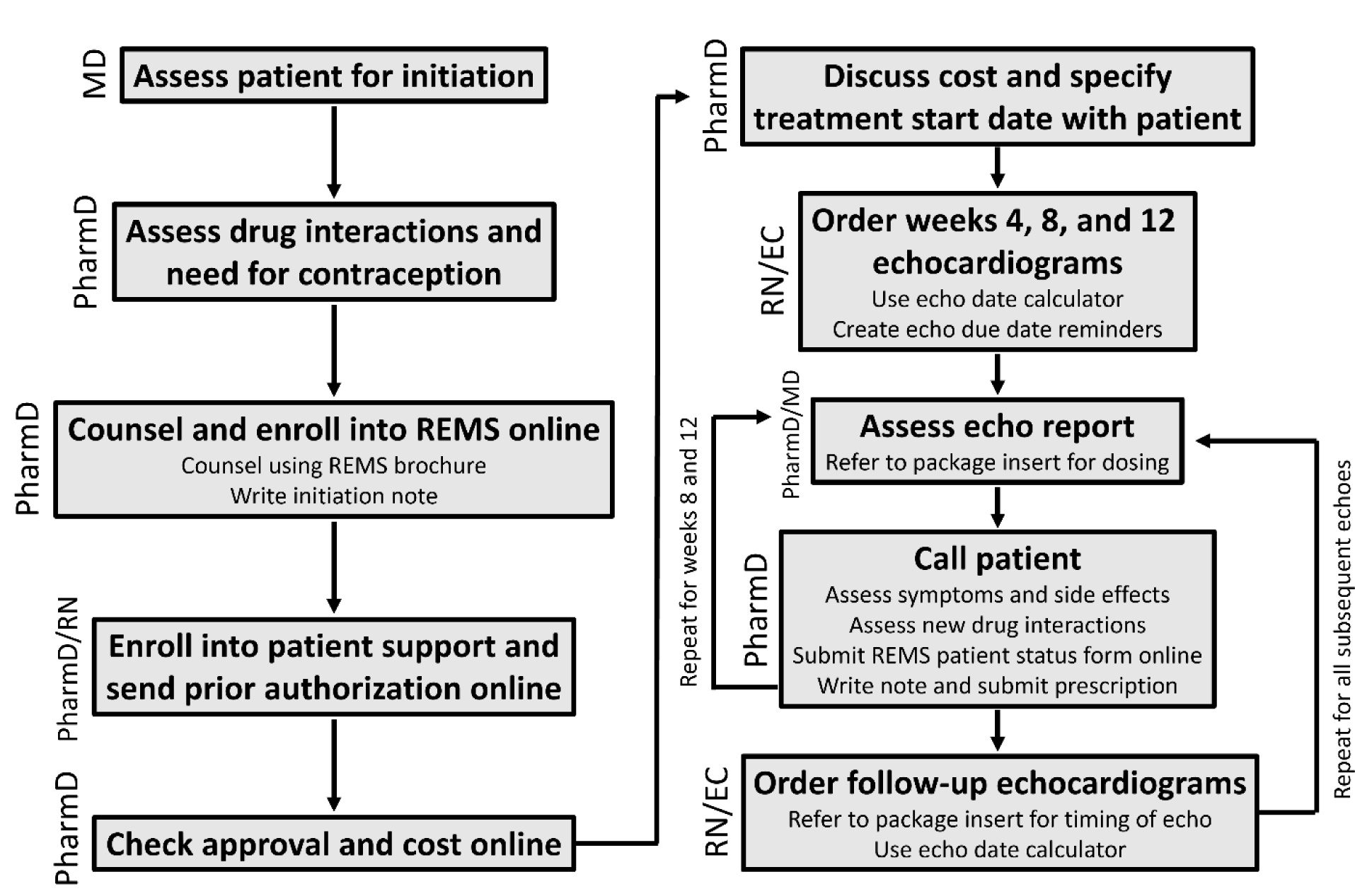
Workflow algorithm. The workflow was used to manage mavacamten for all patients in the study period. Roles for physician (MD), pharmacist (PharmD), nurse (RN), and echo coordinator (EC) are defined. The echo date calculator was created by the authors and calculates expected date ranges for required echocardiograms up to 156 weeks.

Drug interactions are screened using tertiary databases. After counseling using the manufacturer-provided REMS brochure, the pharmacist enrolls the patient into the mavacamten REMS program through the online web portal.^16,17^ A pharmacist note is written in the electronic medical record (EMR) which details New York Heart Association (NYHA) class, baseline echocardiography parameters, medication history, drug interactions, and financial information (Figure S1). To explore cost reduction measures, either the pharmacist or nurse electronically enrolls the patient into the mavacamten patient support program through an online portal.^18^ An electronic prior authorization (PA) and the pharmacist note (Figure S1) are sent to the patient’s insurance through the online portal. After approval and estimated cost are reported in the portal, the pharmacist determines acceptability of cost from the patient and identifies a suitable therapy start date. Follow-up echocardiogram date ranges are determined by entering the start date into a calculator created by the authors which incorporates requirements from the REMS program (File S1). The nurse and echocardiography coordinator schedule the first three REMS-mandated echocardiograms (i.e., weeks 4, 8, and 12), and the start date and scheduled echocardiogram dates are recorded in the EMR. Follow-up echocardiograms are assessed by a cardiologist and a finalized report detailing parameters including LVEF, LVOT, and VLVOT is recorded in the EMR. Necessary dose adjustments based on package insert recommendations are made by the pharmacist and/or cardiologist after assessment of patient symptoms, side effects, and presence of new drug interactions.^19^ REMS patient status forms are submitted by the pharmacist who also writes a note that details the results of the echocardiogram, presence of new drug interactions, and any dose modifications (Figure S2). All follow-up echocardiograms beyond week 12 are managed in a similar manner as described above.

### Data collection

Data for the study was collected through review of the EMR. Baseline characteristics included patient demographics, insurance, vitals, medical and surgical history, NYHA class, LVEF, LVOT, VLVOT, and medications used to treat oHCM such as beta blockers, calcium channel blockers, and disopyramide. Outcomes of interest were the number of patients initiated on therapy, estimated monthly copays before and after exploring cost reduction, time from referral to prior authorization approval, time from referral to first fill, and proportion of days covered (PDC). Time of referral was defined as the point at which the cardiologist referred the patient to the pharmacist for initiation of mavacamten. The PDC is a measure of medication adherence and was calculated as the number of days a patient had mavacamten in hand divided by the number of days between the designated start date and end of study period (May 31, 2023) or therapy discontinuation.^20,21^ Mavacamten dispense dates used in calculating PDC were collected from the online REMS portal. Other outcomes included the change in LVEF, LVOT, and VLVOT in patients who started mavacamten and completed echocardiograms up to 12 weeks or the end of the study period.

### Echocardiography

Patients undergoing echocardiograms were imaged in the left lateral decubitus position, sitting upright with patient legs extended on imaging bed and standing. Imaging was performed using commercially available ultrasound equipment (Philips Epiq CVx Ultrasound, Philips Medical Systems, Andover, MA, USA) and a x5-1 phased array transducer. Before acquisition for ejection fraction, gain, compress and time-compensation were adjusted for optimal endocardial border definition. 2-D Imaging windows included for ejection fraction assessment were the apical 4 and 2 chamber views. Care was taken to include the entire left ventricle while maintaining the highest possible frame rate. Simpson’s Biplane was used to measure the left ventricle during systole and diastole. Resting LVOT peak velocity was measured with Continuous-wave Doppler echocardiography. Color flow Doppler was also placed in the LVOT area to act as a guide for aliasing velocities. Valsalva LVOT interrogation began with patient instructions to exhale forcefully and hold respirations. Continuous wave Doppler imaging and Color Doppler were then activated to obtain outflow velocities. Both resting and Valsalva attempts were made in the 3 and 5 chamber apical views in the three postural positions described above. The highest peak velocities were reported.

### Statistical Analysis

Baseline characteristics are presented as descriptive statistics such as mean (standard deviation [SD]), median (interquartile range [IQR]), or counts (%), where appropriate. Continuous variables were analyzed using two independent samples: the t-test and the Mann-Whitney U test, as appropriate. Categorical variables were analyzed using the chi-square test or Fisher exact test, as appropriate. Generalized estimating equation (GEE) models were used to examine the average change in LVEF, LVOT, VLVOT within 12 weeks of initiating mavacamten, adjusting for age, sex, and ethnicity, and correcting for correlations of repeated measures. The GEE models were limited to 12 weeks given all patients had completed follow-up echocardiograms up to at least this timepoint. All hypotheses were two-sided, and a P-value of <0.05 was considered statistically significant. All statistical analyses were conducted using R statistical software version 4.3.1.

## RESULTS

From May 1, 2022, to May 31, 2023, a total of 34 patients with a history of oHCM were considered for mavacamten initiation. The median age of the study population was 68 years (range 19-88 years), and 17 patients (50%) were female. A total of 19 patients (56%) had a past medical history of hypertension and 5 (15%) had atrial fibrillation. Thirty-two patients (94%) had NYHA class II-III symptoms. The median LVEF was 73.5% (IQR 66 to 77). Median resting LVOT and VLVOT were 37 mmHg (IQR 17 to 64) and 60 mmHg (IQR 45 to 113), respectively. The study patients were on diverse medication regimens. 13 patients (38%) were on disopyramide and beta-blocker, 8 (24%) were on a beta-blocker alone, 4 (12%) were on calcium channel blocker and disopyramide, 4 (12%) were on calcium channel blocker alone, 4 (12%) were not on any medications for oHCM, and 1 (3%) was on beta-blocker and calcium channel blocker. Additional baseline characteristics are detailed in table 1.

**Table 1.** Baseline characteristics. Data are presented as median, number, percentage, or interquartile range. Abbreviations: IQR, interquartile range; LVEF, left ventricular ejection fraction; LVOT, resting left ventricular outflow tract gradient; NYHA, New York Heart Association; VLVOT, Valsalva left ventricular outflow tract gradient.

| **Table 1. Baseline characteristics** |  |  |
| --- | --- | --- |
| **Characteristic** | **All patients considered for therapy (n = 34)** | **Patients initiated on therapy (n = 21)** |
| Age, years (range) | 68 (19-88) | 64 (48, 70) |
| Female, n (%) | 17 (50) | 11 (52) |
| Body-mass index, kg/m<sup>2</sup> (IQR) | 27.7 (24.4, 30.4) | 28.7 (24.9, 31.0) |
| Systolic blood pressure, mmHg (IQR) | 130 (116, 141) | 126 (115.5, 142) |
| Heart rate, beats per minute (IQR) | 66 (60,75) | 65 (59, 74) |
| Ethnicity, n (%) |  |  |
| White | 20 (59) | 11 (52) |
| Hispanic | 7 (21) | 4 (19) |
| Asian | 4 (12) | 3 (14) |
| Unidentified | 3 (9) | 3 (14) |
| Medical history, n (%) |  |  |
| Hypertension | 19 (56) | 11 (52) |
| Atrial fibrillation | 5 (15) | 1 (5) |
| Surgical history, n (%) |  |  |
| Septal ablation | 4 (12) | 3 (14) |
| Myectomy | 2 (6) | 2 (10) |
| NYHA Class, n (%) |  |  |
| Class II | 20 (59) | 2 (10) |
| Class III | 12 (35) | 10 (48) |
| Class IV | 2 (6) | 9 (43) |
| Echocardiographic parameters |  |  |
| LVEF, % (IQR) | 73.5 (66, 77) | 75 (69, 80) |
| LVOT, mmHg (IQR) | 37 (17, 64) | 49 (19.5, 75) |
| VLVOT, mmHg (IQR) | 60 (45, 113) | 73 (52, 125.5) |
| Background therapy, n (%) |  |  |
| Disopyramide + beta-blocker | 13 (38) | 7 (33) |
| Beta-blocker | 8 (24) | 5 (24) |
| Disopyramide + calcium channel blocker | 4 (12) | 3 (14) |
| Calcium channel blocker | 4 (12) | 2 (10) |
| Calcium channel blocker + beta-blocker | 1 (3) | 1 (5) |
| No medication | 4 (12) | 3 (14) |
| Insurance coverage, n (%) |  |  |
| Medicare | 12 (35) | 5 (24) |
| Commercial | 11 (32) | 9 (43) |
| Medicaid | 7 (21) | 6 (29) |
| Medicare-Medicaid | 2 (6) | 1 (5) |
| Military | 1 (3) | 0 (0) |
| None | 1 (3) | 0 (0) |

Out of 34 patients considered for treatment with mavacamten, 21 patients (62%) were successfully initiated on treatment. The reasons for non-initiation varied. 5 patients (15%) declined due to the medication cost, 5 (15%) declined due to echocardiogram requirements, 1 (3%) was not initiated due to a drug interaction, and 1 (3%) did not start therapy due to the medication not being included on their insurance formulary. Additionally, 1 patient was in the process of awaiting medication approval at the end of the study period.

Figure 2 illustrates the cost outcomes by insurance type for 28 patients that had cost assessment completed, with individual lines indicating the monthly co-pays. Six patients did not undergo cost assessment due to declining echocardiogram follow-up or therapy not being on formulary. The ‘Pre-Out of Pocket Cost’ represents each patient’s initial cost per CoverMyMeds before any cost reduction strategies, such as copay cards or patient assistance programs. ‘Post-Out of Pocket Cost’ reflects the actual cost of medication after employing these reduction strategies. For the Medicare cohort (n = 10), the median pre-out of pocket monthly co-pay was $1,801, ranging from $79.99 to $3,856. Following cost reduction efforts, the median post-out of pocket monthly cost expense decreased to $79, with a range from $0 to $3,856. In the group with commercial insurance (n = 9), the median pre-out of pocket monthly cost stood at $120, with individual costs ranging between $25 and $2,850. After applying cost reductions, the post-out of pocket monthly cost uniformly dropped to $10. All patients with Medicaid (n = 7) and those with combined Medicare-Medicaid coverage (n = 2) were not eligible for cost reduction strategies. For these patients, copays ranged between $0 and $4.

**Figure 2.**
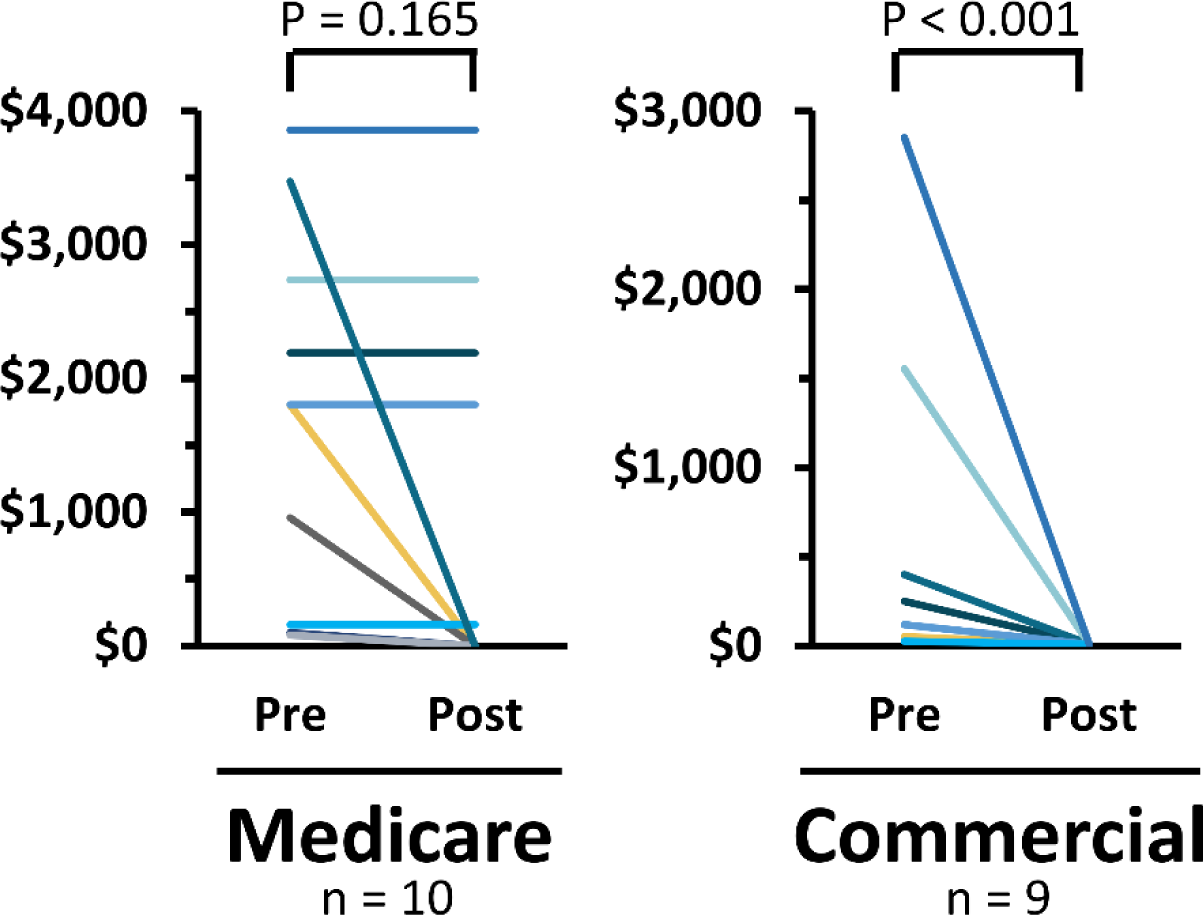
Cost outcomes. Lines indicate monthly copays for individual patients. Pre = cost per CoverMyMeds before exploring cost reduction (e.g., copay cards and patient assistance). Post = actual cost after reduction methods were explored. All Medicaid (n = 7) and Medicare/Medicaid (n = 2) copays were not eligible for cost reduction but ranged between $0 and $4. Median cost is compared. A P value <0.05 indicates significance.

Of the 34 patients, 19 had prior authorization completed. The median time to approval was 5 days (IQR 12 days, 1-13 days). Time from initial referral to first fill for the 21 patients initiated on mavacamten was a median 22 days (IQR 25 days, 15-40 days). The average PDC for these patients was 99.1%.

Figure 3 shows individual trends in LVEF, LVOT, and VLVOT for 18 of the 21 patients who started mavacamten through the end of the study period (May 31, 2023). Three patients are not shown given they discontinued therapy within 4 weeks of starting therapy. One patient discontinued due to worsening heart failure after stopping disopyramide for mavacamten. This case is described elsewhere.^22^ The second patient stopped therapy due to progression of her chronic kidney disease and placement on permanent renal replacement therapy, an area of care that is lacking data for mavacamten efficacy and safety. The third patient stopped therapy due to intolerable dizziness. Overall, no patients experienced a decrease in LVEF to less than 50%. All patients received labeled dose changes to mavacamten except for patient 12. This patient had a dose reduction from 10 mg to 5 mg daily at week 36 due to concern for impending systolic dysfunction after observing a 17-percentage point reduction in LVEF from 73% to 56% between weeks 24 and 36 of therapy. At week 40 while taking the reduced 5 mg daily, the patient’s LVEF, LVOT, and VLVOT were 68%, 120 mmHg, and 131 mmHg, respectively. After further review of echocardiograms, the decrease in LVEF between weeks 24 and 36 were thought secondary to variability in endocardial boarder tracing during biplane echocardiography. Mavacamten was subsequently increased back to 10 mg daily.

**Figure 3.**
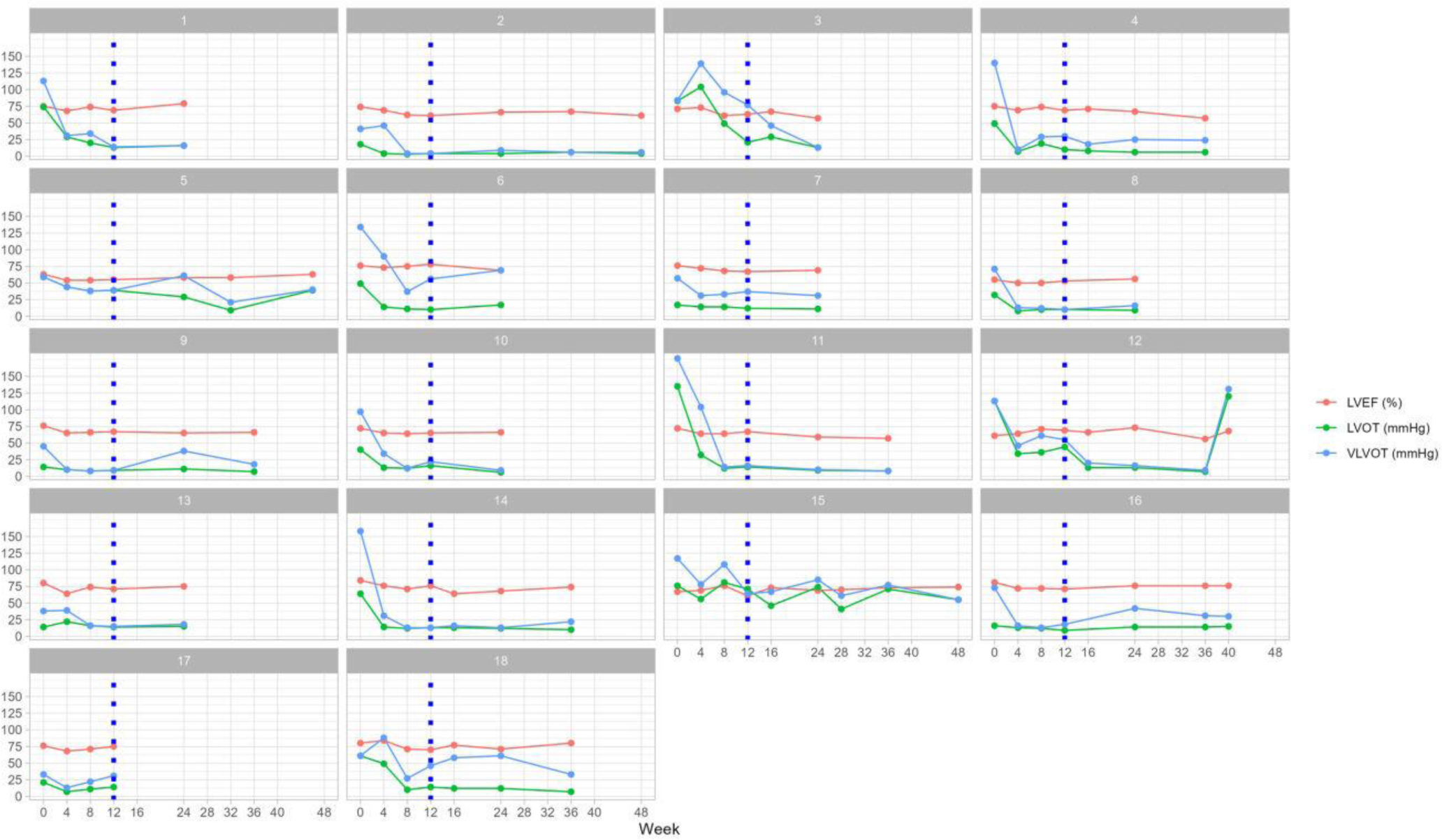
Individual trends in echocardiogram parameters. Trends in LVEF, LVOT, and VLVOT for 18 of the 21 patients started on mavacamten are shown. Data are incomplete given patients were initiated at different times throughout the study period and then followed until the end of study. The blue dotted line designates the end of the initial 12-week phase of mavacamten therapy. Dose changes were made in accordance with labeled recommendations. Echocardiograms per the Risk Evaluation and Mitigation Strategy program were completed at each dotted timepoint. Abbreviations: LVEF, left ventricular ejection fraction; LVOT, resting left ventricular outflow tract gradient; VLVOT, Valsalva left ventricular.

Figure 4 shows the effect of mavacamten 5 mg daily on LVEF, LVOT, and VLVOT across the 12-week initiation phase. The results from the GEE model for the change in LVEF, LVOT, and VLVOT are shown in Table 2. There was a significant change in all measures across time within 12 weeks from initiating mavacamten. VLVOT had the highest average reduction (−4.79 mmHg; 95% CI = −0.75, 0.15) per week, followed by LVOT (−2.63; 95% CI = −3.89, 1.37), and LVEF (−0.45 mmHg; 95% CI = −0.75, −0.15) respectively. Interestingly, ethnicity was a significant predictor of resting LVOT and VLVOT. On average, Hispanic patients had a lower LVOT and VLVOT by 21.9 (95% CI = −32.5, −11.3) and 30.7 (95% CI = −45.3, −16.1) mmHg when compared to White patients. Age and sex were predictors of LVEF only. For every 10 years, LVEF increases by 0.930% (95% CI = 0.103, 1.76). Male patients had a lower LVEF by 5.67% (95% CI = −8.29, −3.05).

**Figure 4.**
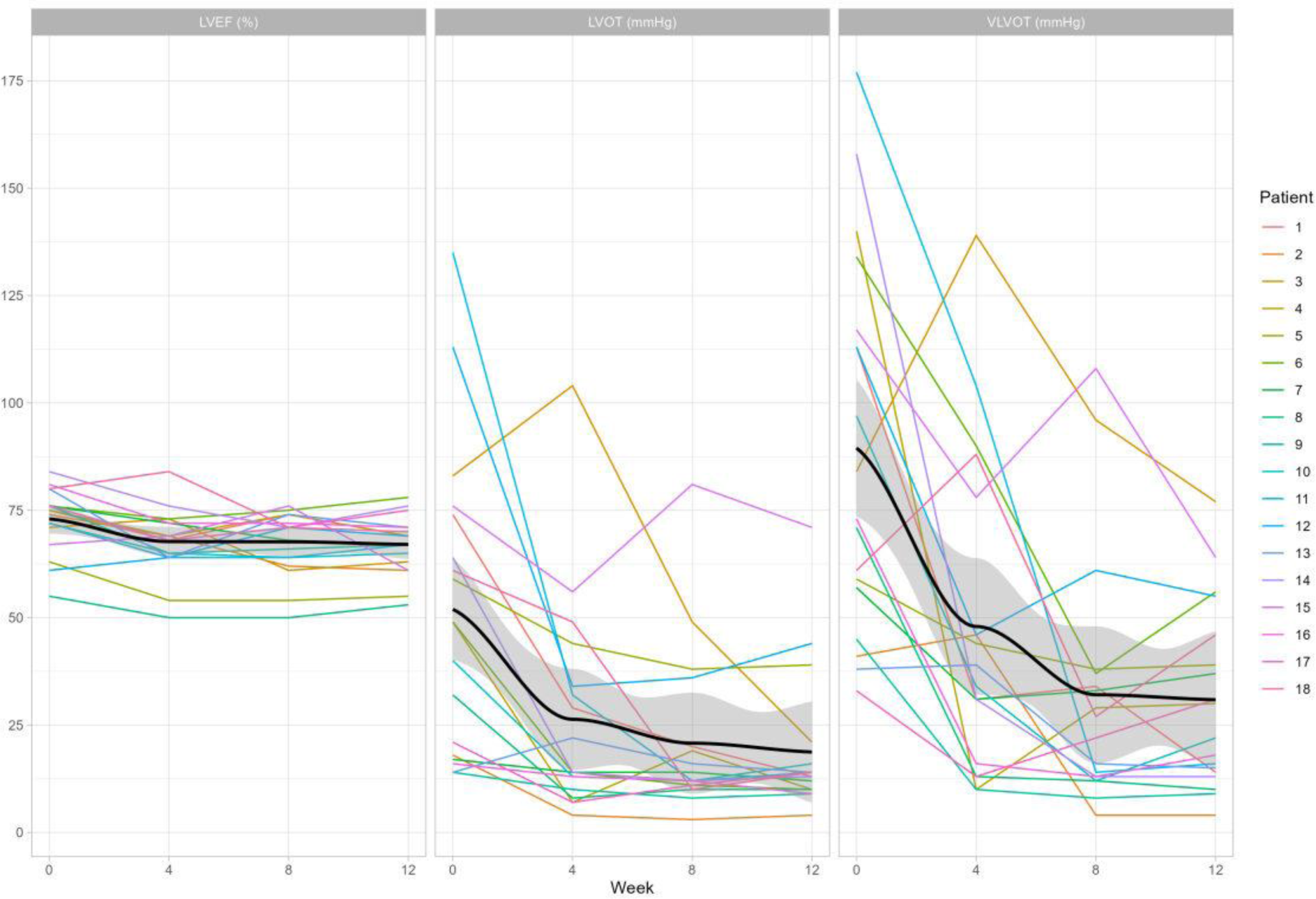
Effect of mavacamten on echocardiogram parameters over 12 weeks. Spaghetti plots showing the change in LVEF, LVOT, and VLVOT are shown. Each colored line represents an individual patient. All patients were treated with 5 mg daily mavacamten over the 12-week period. The black line indicates the smoothed average. Gray area represents the 95% confidence interval. Abbreviations: LVEF, left ventricular ejection fraction; LVOT, resting left ventricular outflow tract gradient; VLVOT, Valsalva left ventricular.

**Table 2.** Generalized estimating equation model for the change in echocardiographic parameters over 12 weeks of mavacamten therapy. Abbreviations: CI, confidence interval; LVEF, left ventricular ejection fraction; LVOT, resting left ventricular outflow tract gradient; VLVOT, Valsalva left ventricular outflow tract gradient. A P value <0.05 indicates significance.

| **Table 2. Generalized estimating equation model for the change in echocardiographic parameters over 12 weeks of mavacamten therapy** |  |  |  |  |  |  |  |  |  |
| --- | --- | --- | --- | --- | --- | --- | --- | --- | --- |
|  | **LVEF** |  |  | **LVOT** |  |  | **VLVOT** |  |  |
|  | Estimate | 95% CI | P-Value | Estimate | 95% CI | P-Value | Estimate | 95% CI | P-Value |
| Age, 10 years | 0.930 | 0.103, 1.756 | 0.027 | -2.40 | -5.52, 0.719 | 0.13 | -0.434 | -4.42, 3.56 | 0.83 |
| **Sex** |  |  |  |  |  |  |  |  |  |
| Female | Ref |  |  | Ref |  |  | Ref |  |  |
| Male | -5.67 | -8.29, -3.05 | < 0.001 | 7.09 | -3.33, 17.5 | 0.18 | 2.167 | -12.3, 16.6 | 0.77 |
| **Ethnicity** |  |  |  |  |  |  |  |  |  |
| White | Ref |  |  | Ref |  |  | Ref |  |  |
| Asian | 0.994 | -2.67, 4.65 | 0.60 | 14.8 | -5.48, 35.1 | 0.153 | 18.1 | -15.1, 51.2 | 0.29 |
| Hispanic | -2.49 | -6.84, 1.85 | 0.26 | -21.9 | -32.5, -11.3 | < 0.001 | -30.7 | -45.3, -16.1 | <0.001 |
| Unidentified | 1.67 | -1.77, 5.11 | 0.34 | -11.4 | -24.7, 1.85 | 0.09 | -25.3 | -42.4, -8.28 | 0.007 |
| Time, per week | -0.45 | -0.75, -0.15 | 0.004 | -2.63 | -3.89, 1.37 | < 0.001 | -4.79 | -6.42, -3.17 | <0.001 |

## DISCUSSION

Management of mavacamten is complex, requiring frequent echocardiogram monitoring, drug interaction screening, and effort to lower patient-level costs. We created a workflow to streamline mavacamten management by designating roles for clinic personnel, developing tools for record keeping (i.e., the date calculator and EMR reminder lists), and leveraging available resources from the manufacturer’s patient support program. This workflow was successfully implemented in our clinic and effectively addressed many of the challenges of the medication.

We initiated mavacamten in 2 patients with NYHA class IV symptoms, a criterion for exclusion in EXPLORER-HCM.^13^ The 2 patients with class IV symptoms had baseline VLVOT gradients of 113 and 59 mmHg while on disopyramide and a beta blocker. Although they met criteria and were offered SRT, they did not want to pursue intervention and instead tried mavacamten.^23^ Currently, these 2 patients have been on mavacamten for more than a year, report improved symptoms, and have VLVOT gradients of less than 50 mmHg which makes them no longer eligible for SRT. While mavacamten is not indicated in NYHA class IV oHCM, the phase three VALOR-HCM study established efficacy and safety of the medication in this patient population.^14^ Our experience further demonstrates feasibility of starting mavacamten in real-world patients with NYHA class IV symptoms.

Half of our patients were on disopyramide at baseline and were considered for transition to mavacamten to ameliorate disopyramide-related anticholinergic side effects and multiple times a day dosing. While VALOR-HCM included patients on disopyramide, we did not continue disopyramide when starting mavacamten.^14^ This is in accordance with labeled recommendations to avoid concomitant use of the therapies out of concern for excessive additive negative inotropy and insufficient data to demonstrate safety of co-administration.^19^ We tapered off disopyramide when starting mavacamten following a published guide that incorporates dose, frequency, and half-life of the two therapies.^22^ This method of tapering disopyramide allows for management of obstructive symptoms while minimizing the risk for adverse effects or worsening left ventricular dysfunction when starting mavacamten.

We identified Hispanic ethnicity as a predictor of LVOT and VLVOT. This observation is most likely attributed to the inherent sampling variability associated with a small sample size.^24^ It is worth noting that Mavacamten metabolism is decreased in poor CYP 2C19 metabolizers; however, this phenotype is not appreciably frequent in Hispanic populations.^15,25,26^ Overall, this finding should be interpreted with caution, given this study was not designed to investigate the effects of ethnicity on mavacamten response.

Cost of therapy is an important consideration that is a challenge for patients. A published pharmacoeconomic analysis showed that mavacamten produces more quality-adjusted life-years than beta blocker, calcium channel blocker, and disopyramide therapy but at much higher additional costs.^27^ The yearly price of mavacamten based on its wholesale acquisition cost is $89,500, equating to $7,458 a month for patients without insurance.^28^ To improve affordability, we enrolled all patients into the manufacturer’s support program regardless of their insurance type which resulted in greatly lowered monthly out-of-pocket costs. Half of the patients with Medicare Part D insurance were provided third party grant funding which lowered copays to $0 a month. Furthermore, all patients with commercial insurance had copays lowered to $10 a month after leveraging assistance from the support program.

Time to PA approval was highly dependent on the response rate from the payor, but we achieved a median time to PA approval of 5 days which is likely driven by use of the online portal and preemptively uploading the pharmacist initiation note during the application process. The time to first fill was a median 22 days and was highly driven by patient choice. Multiple patients elected to delay their start date and, thus, the first dispense date of medication.

The PDC is a measure of adherence endorsed by multiple pharmacy quality assurance organizations and has a standard adherence threshold of 80%.^20,21,29^ The average PDC for patients initiated on therapy in this study was 99.1%. This high level of adherence is likely influenced by the requirement for REMS-mandated echocardiograms. Under the program, patients are unable to get medication without complete echocardiogram follow-up. This requirement was heavily stressed during counseling prior to initiation which resulted in 5 patients declining therapy due to unwillingness to follow-up for echocardiograms. Regardless, all 21 accepting patients started on therapy had excellent adherence which may be reflective of the high degree of patient engagement involved in the workflow.

The described workflow has specific characteristics that are key to its efficiency. First, the initial 3 echocardiograms are scheduled immediately after starting mavacamten which allows for appropriate advanced planning for the patient. Furthermore, all echocardiogram date ranges are determined using a calculator that calculates dates out to 156 weeks. The online calculator provided by the manufacturer is limited to the first 12 weeks of therapy.^30^ Second, follow-up with patients is completed primarily over the phone. While assessment of symptoms, new drug interactions, and counseling on dose changes is ideally performed in-person, high volume centers may have difficulty scheduling patients with the cardiologist particularly during the first 12 weeks of therapy when echocardiograms are completed monthly. Our workflow has patients follow up in-person with the cardiologist according to provider availability while REMS echocardiograms are followed up over the phone. Third, all forms (i.e., REMS, patient support, and PA) are submitted online. This mode of communication improves information delivery by eliminating the reliance on faxing and instances where completed paper forms may be lost.

The workflow has specific limitations. The effectiveness of the workflow is in part a result of the distribution of responsibilities among the team members. We acknowledge that many centers do not have a team of physicians, pharmacists, nurses, and echocardiography coordinators to manage mavacamten. We recommend designating suitable clinic personnel to the steps described in Figure 1 to create an individualized workflow for these centers. While our patient cohort was limited to 21 patients on therapy, centers with larger cohorts can still use the described workflow given the steps are uniformly required, except for patient access investigation, by the REMS program. Effectiveness with respect to lowering copays in our workflow is dependent on patient support programs which are theorized to increase payor costs by undercutting the tiered formulary system, ultimately leading to increased patient costs through raised premiums.^31,32^ It is important to consider overall societal cost and, thus, we recommend initiating mavacamten in patients who are intolerant to or are still symptomatic while on current guideline therapy. While payor costs will be increased, use of mavacamten in patients without prior history of therapy may also be reasonable given there is a lack of robust clinical data supporting the use of beta blockers, calcium channel blockers, and disopyramide in oHCM.^33^ Future studies comparing mavacamten to these alternatives are needed.

## CONCLUSION

The multidisciplinary workflow described here addressed the challenges associated with real-world mavacamten management by designating roles for clinic personnel, implementing tools for record-keeping, and leveraging manufacturer patient support programs. Given the workflow has institution-specific characteristics contributing to its effectiveness, we acknowledge the need for adaptation in centers with different resources. Regardless, this report provides valuable insights for other clinics aiming to implement a structured workflow for mavacamten management.

## Data Availability

Data presented are not openly available due to institutional policy.

## Non-standard Abbreviations and Acronyms

CYP: cytochrome P450
EMR: electronic medical record
GEE: generalized estimating equation
HCM: hypertrophic cardiomyopathy
LVEF: left ventricular ejection fraction
LVOT: left ventricular outflow tract
NYHA: New York Heart Association
oHCM: obstructive hypertrophic cardiomyopathy
PA: prior authorization
PDC: proportion of days covered
REM: Risk Evaluation and Mitigation Strategy
SRT: septal reduction therapy
VLVOT: Valsalva left ventricular outflow tract

## SOURCES OF FUNDING

This study was unfunded.

## DISCLOSURES

AW serves as a clinical consultant for Bristol Myers Squibb and Cytokinetics. JSE reports receiving honoraria from Bristol Myers Squibb. ZY serves as a clinical consultant for Takeda Pharmaceuticals. HJ, TH, MK, MR, and TB have no conflicts of interest to disclose.

## Notes

### Clinical Trial

This is retrospective study. A clinical trial ID was not required.

### Author Declarations

This study was approved by the UC San Diego Health institutional review board. Informed consent was not required.

